# Test-retest reliability of functional connectivity in depressed adolescents

**DOI:** 10.1101/2022.10.11.22280962

**Authors:** Chris C. Camp, Stephanie Noble, Dustin Scheinost, Argyris Stringaris, Dylan M. Nielson

## Abstract

The test-retest reliability of fMRI functional connectivity is a key factor in the identification of reproducible biomarkers for psychiatric illness. Low reliability limits the observable effect size of brain-behavior associations. Despite this important connection to clinical applications of fMRI, few studies have explored reliability in populations with psychiatric illnesses or across age groups. We investigate the test-retest reliability of functional connectivity in a longitudinal cohort of adolescents with and without major depressive disorder (MDD). Measuring reliability is complex and several metrics exist that can offer unique perspectives: for example, univariate metrics capture reliability of a single connection at a time while multivariate metrics reflect stability of the entire connectome. We compare a widely used univariate metric, intraclass correlation coefficient (ICC), and two multivariate metrics, fingerprinting and discriminability. Depressed adolescents were more reliable than healthy adolescents at the univariate level (0.34 > 0.24; Wilcoxon rank-sum: *p* < .001), and both groups had poor average ICCs (<0.4). Multivariate reliability was high in both groups: fingerprinting (FI_HV_ = 0.53; FI_MDD_ = 0.45; Poisson(1) test *p* < .001) and discriminability were above chance (*Discr*_HV_ = 0.75; *Discr*_MDD_ = 0.76; 500-fold permutation test *p* < .01). Reliability was not associated with symptoms or medication, suggesting that there is not a strong relationship between depression and reliability. These findings support the shift towards multivariate analysis for improved power and reliability.

## INTRODUCTION

One of the foremost goals of neuroimaging work in psychiatry is to elucidate the brain correlates of psychiatric illnesses. However, a major barrier to identifying these is the reproducibility of neuroimaging findings. The “replication crisis” has highlighted the difficulty of reproducing underpowered neuroimaging results, especially from functional magnetic resonance imaging (fMRI). This lack of power is inseparable from the reliability of the data. Reliability places an upper bound on any observable effect size, and in turn limits statistical power (Zuo et al., 2019). Thus, quantifying the “test-retest reliability”—that is, the reliability of a test or measurement over repetitions—is critical to interpreting the validity of results.

Nevertheless, quantifying the reliability of fMRI is complex with many factors influencing it. Longer scans and shorter intervals between scans increase reliability, while artifact correction can decrease it—underscoring the separation between validity and reliability, as noise such as motion can be highly reliable (Noble et al., 2021). Validity, the accuracy with which a measure represents the “ground truth” of a desired construct, is impossible to fully determine in fMRI due to the thousands of complex interactions that produce the blood oxygenation-level dependent (BOLD) signal. However, measures must be reliable to be valid, highlighting the importance of these investigations. The measurement of test-retest reliability can be done through different metrics, each of which reflect unique forms of reliability subject to interpretation. These metrics can be univariate—reflecting the reliability of each test item or measurement individually—or multivariate—reflecting the stability of multidimensional data, such as whole-brain patterns.

Univariate measures, including the widely used intraclass correlation coefficient (ICC), are typically poor in fMRI (Elliott et al., 2020; Noble et al., 2017, 2019). In contrast, functional connectivity – the correlations between regional activity – has high multivariate reliability (E. W. Bridgeford et al., 2021; Horien et al., 2019; Noble et al., 2017), putatively because multivariate approaches incorporate higher dimensionality variance structure. Consequently, there have been several efforts to understand the nuance of reliability and its interpretation to optimize our data collection and processing methods.

Despite these effects and the larger goals of neuroimaging in psychiatric research, few works have investigated test-retest reliability of functional connectivity in a psychiatric population and compared it to a similar healthy population. Those that have were focused on adult populations with either mild cognitive impairment (Blautzik et al., 2013) or schizophrenia (Manoach et al., 2001). Assessing reliability in psychiatric populations is necessary for guiding the search for brain-behavior associations that can predict, diagnose, or explain illnesses (Zuo & Xing, 2014).

We investigated the test-retest reliability of resting state functional connectivity in a cohort of adolescents with and without major depressive disorder. Major depressive disorder is the leading cause of disability worldwide and has proven one of the most challenging illnesses for reproducible biomarker identification (Fried et al., 2022; Nielson et al., 2020; WHO, 2017).

Depression typically develops during adolescence, marking this a critical time for investigating neural changes that could indicate depression onset. By characterizing the stability of connectomes over a one-year period, we can begin to understand how age and psychiatric illness might affect reliability, informing clinical applications of fMRI. For this reason, we focus on resting state functional connectivity, which is measured while a participant lays at rest in a scanner. This approach is frequently used for biomarker identification due to its accessibility for an array of clinical populations (Nour et al., 2022). We employ univariate ICC and two multivariate measures of reliability: fingerprinting (Finn et al., 2015) and discriminability (E. W. Bridgeford et al., 2021). Functional connectome fingerprinting reflects the proportion of subjects whose connectomes are most correlated with their own at a later timepoint. High fingerprinting accuracy has been observed in several datasets, suggesting that functional connectivity data is stable and unique enough to reliably identify subjects (Horien et al., 2019). Discriminability is a multivariate reliability metric that is robust to noise and provides an upper bound on classification accuracy (E. W. Bridgeford et al., 2021). Initial results suggest that functional connectivity data are highly discriminable. We thus combined ICC, fingerprinting, and discriminability to determine how these measures may reflect different facets of reliability and offer unique perspectives on the data. In conducting these analyses, we expected adolescents with depression to have less reliable connectomes than their healthy peers, in line with previous work on other psychiatric illness (Blautzik et al., 2013; Manoach et al., 2001). We also hypothesized that multivariate reliability (fingerprinting and discriminability) would be higher than univariate (ICC). Through this investigation, we will clarify the test-retest reliability of functional connectivity in a clinically relevant population, guiding the search for biomarkers that can revolutionize psychiatry.

## METHODS

### Participants

Participants were part of the National Institute of Mental Health Characterization and Treatment of Depression (NIMH CAT-D) cohort, a longitudinal case-control study. Adolescent volunteers (age 12–19 years) were recruited through mail, online advertisement and direct referrals from clinical sources. Participants provided informed consent to a protocol approved by the NIH Institutional Review Board (clinical trial no. NCT03388606) before completing questionnaires and an in-person evaluation with a medical practitioner at the NIH clinical center to guarantee their suitability to enroll in the study. Both healthy volunteers (not satisfying criteria for any diagnosis according to DSM-5) and patients with a primary diagnosis of major depression (MDD) or sub-threshold depression were included. After one year, participants returned for a follow-up visit consisting of questionnaires, a clinical interview, and an additional scan.

### Clinical interview and questionnaires

Participants were diagnosed through the Kiddie-Schedule for Affective Disorders and Schizophrenia (K-SADS), a semi-structured interview (Kaufman & Schweder, 2004). Questionnaires administered included the Child Self-Report: Short Version Mood and Feelings Questionnaire (S-MFQ; Angold et al., 1995), Affective Reactivity Index – 1 week (ARI; Stringaris et al., 2012), Snaith-Hamilton Pleasure Scale (SHAPS; Snaith et al., 1995), and Screen for Child Anxiety Related Disorders (SCARED; Birmaher et al., 1997).

### fMRI data acquisition

Following in-person screening, participants were scanned in a General Electric (Waukesha, WI) Signa 3-Tesla MR-750s magnet with 32 channel head coils, being randomly assigned to one of two similar scanners. Both scanners were housed in the NMR suite of the NIH clinical center. The fixation stimulus was displayed via back-projection from a head-coil-mounted mirror. Foam padding was used to constrain head movement. Resting state data was collected with a multi-echo T2*-weighted echo-planar sequence with 32 oblique axial slices (4.0 mm thickness) (4 echos: 15.2 ms, 28.8 ms, 42.4 ms, 56 ms; flip angle, 80°; 64 × 64 matrix; field of view, 240 mm; in-plane resolution, 3.75 mm × 3.75 mm; repetition time was 2500 ms). To improve the localization of activations, a high-resolution structural image was also collected from each participant during the same scanning session using a T1-weighted standardized magnetization prepared spoiled gradient recalled echo sequence with the following parameters: 176 1 mm sagital slices; repetition time, 7.7 ms; echo time, 3.436 ms; flip angle, 7°; 256 × 256 matrix; field of view, 256 mm; in-plane resolution, 1.0 mm × 1.0 mm. During this structural scanning session, all participants watched a short neutral-mood documentary movie about bird migration.

We collected fMRI data from 202 volunteers that passed our inclusion criteria. Of this sample, 10 participants were excluded from all reported analysis due to issues with data collection, quality, or processing. To have a consistent time interval between participants, we only included baseline and 1-year sessions and only included participants who had usable data from both sessions, excluding 104 participants. This left 88 participants (57 MDD; 64 females; median age at baseline: 15.71).

### Data preprocessing

Results included in this manuscript come from preprocessing performed using fMRIPrep 21.0.0 (Esteban et al., 2019; RRID:SCR_016216), which is based on Nipype 1.6.1 (Esteban, Oscar et al., 2022; Gorgolewski et al., 2011; RRID:SCR_002502).

#### Preprocessing of B0 inhomogeneity mappings

A total of 2 fieldmaps for the resting state scans were collected for each subject. A B0-nonuniformity map (or fieldmap) was estimated based on two (or more) echo-planar imaging (EPI) references with topup (Andersson et al., 2003; FSL 6.0.5.1:57b01774).

#### Anatomical data preprocessing

A T1-weighted (T1w) images was collected for each session. All of them were corrected for intensity non-uniformity (INU) with N4BiasFieldCorrection (Tustison et al., 2010), distributed with ANTs 2.3.3 (Avants et al., 2008, RRID:SCR_004757). The T1w-reference was then skull-stripped with a Nipype implementation of the antsBrainExtraction.sh workflow (from ANTs), using OASIS30ANTs as target template. Brain tissue segmentation of cerebrospinal fluid (CSF), white-matter (WM) and gray-matter (GM) was performed on the brain-extracted T1w using fast (FSL 6.0.5.1:57b01774, RRID:SCR_002823, Zhang et al., 2001). A T1w-reference map for each subject was computed after registration of all of the T1w images (after INU-correction) using mri_robust_template (FreeSurfer 6.0.1, Reuter et al., 2010). Brain surfaces were reconstructed using recon-all (FreeSurfer 6.0.1, RRID:SCR_001847, Dale et al., 1999), and the brain mask estimated previously was refined with a custom variation of the method to reconcile ANTs-derived and FreeSurfer-derived segmentations of the cortical gray-matter of Mindboggle (RRID:SCR_002438, Klein et al., 2017). Volume-based spatial normalization to MNI152NLin2009cAsym was performed through nonlinear registration with antsRegistration (ANTs 2.3.3), using brain-extracted versions of both T1w reference and the T1w template. The following templates were selected for spatial normalization: ICBM 152 Nonlinear Asymmetrical template version 2009c [Fonov et al., (2009), RRID:SCR_008796; TemplateFlow ID: MNI152NLin2009cAsym], FSL’s MNI ICBM 152 non-linear 6th Generation Asymmetric Average Brain Stereotaxic Registration Model [Evans et al., (2012), RRID:SCR_002823; TemplateFlow ID: MNI152NLin6Asym].

#### Functional data preprocessing

For each of the resting-state runs found per subject (across all tasks and sessions), the following preprocessing was performed. First, a reference volume and its skull-stripped version were generated from the shortest echo of the BOLD run using a custom methodology of fMRIPrep. Head-motion parameters with respect to the BOLD reference (transformation matrices, and six corresponding rotation and translation parameters) are estimated before any spatiotemporal filtering using mcflirt (FSL 6.0.5.1:57b01774, Jenkinson et al., 2002). The estimated fieldmap was then aligned with rigid-registration to the target EPI (echo-planar imaging) reference run. The field coefficients were mapped on to the reference EPI using the transform. BOLD runs were slice-time corrected to 1.21s (0.5 of slice acquisition range 0s-2.42s) using 3dTshift from AFNI (Cox & Hyde, 1997, RRID:SCR_005927). A T2★ map was estimated from the preprocessed EPI echoes, by voxel-wise fitting the maximal number of echoes with reliable signal in that voxel to a monoexponential signal decay model with nonlinear regression. The T2★/S0 estimates from a log-linear regression fit were used for initial values. The calculated T2★ map was then used to optimally combine preprocessed BOLD across echoes following the method described in (Posse et al., 1999). The optimally combined time series was carried forward as the preprocessed BOLD. The BOLD reference was then co-registered to the T1w reference using bbregister (FreeSurfer) which implements boundary-based registration (Greve & Fischl, 2009). Co-registration was configured with six degrees of freedom. First, a reference volume and its skull-stripped version were generated using a custom methodology of fMRIPrep. Several confounding time-series were calculated based on the preprocessed BOLD: framewise displacement (FD), DVARS and three region-wise global signals. FD was computed using two formulations following Power (absolute sum of relative motions, Power et al. (2014)) and Jenkinson (relative root mean square displacement between affines (Jenkinson et al., 2002). FD and DVARS are calculated for each functional run, both using their implementations in Nipype (following the definitions by Power et al., 2014). Frames that exceeded a threshold of 0.5 mm FD as well as following frames were annotated as motion outliers and censored in all analyses. Additionally, frames in which more than 10% of voxels were temporal outliers were censored in all analyses. The BOLD time-series were resampled into standard space, generating a preprocessed BOLD run in MNI152NLin2009cAsym space. First, a reference volume and its skull-stripped version were generated using a custom methodology of fMRIPrep. All resamplings can be performed with a single interpolation step by composing all the pertinent transformations (i.e. head-motion transform matrices, susceptibility distortion correction when available, and co-registrations to anatomical and output spaces). Gridded (volumetric) resamplings were performed using antsApplyTransforms (ANTs), configured with Lanczos interpolation to minimize the smoothing effects of other kernels (Lanczos, 1964).

Many internal operations of fMRIPrep use Nilearn 0.8.1 (Abraham et al., 2014, RRID:SCR_001362), mostly within the functional processing workflow. For more details of the pipeline, see the section corresponding to workflows in fMRIPrep’s documentation.

#### Copyright Waiver

The above boilerplate text was automatically generated by fMRIPrep with the express intention that users should copy and paste this text into their manuscripts unchanged. It is released under the CC0 license.

### Functional connectivity analysis

Nodes were defined with the 122-cluster Bootstrapped Analysis of Stable Clusters (BASC) atlas (Bellec et al., 2010). Voxel timeseries were averaged within each cluster and the resulting mean timeseries were used to calculate the resting state functional connectivity. We regressed out motion and physiological confounds and included cosine terms to control for temporal drifts. Regressors for heart rate, respiration, and respiration volume per time were created from cardiac and respiratory data with AFNI’s RetroTS.py (Cox, 1996). We included 24 motion terms, translations and rotations in each axis, those values squared, their derivatives, and derivatives squared. Using the Nilearn software package, pairwise correlations were computed between all nodes to generate a 122×122 functional connectivity matrix – also known as a connectome – for each individual (Abraham et al., 2014). These matrices form the basis of the subsequent reliability analysis.

### Intraclass correlation coefficient

To assess univariate reliability of the functional connectomes, ICCs were calculated using the *psych* R package with the healthy and depressed populations (Revelle, 2022). We performed absolute agreement, two-way random effects model reliability assessment, or Shrout and Fleiss Convention ICC(2,1), which models the raters (in this case, scanners) as randomly selected from a larger group (Shrout & Fleiss, 1979). Subjects were bootstrapped 1000 times with replacement to ascertain confidence intervals. ICCs were then averaged across all edges to obtain group means. The criteria for ICC values are typically represented as: poor < 0.4, fair 0.4-0.59, good 0.6-0.74, excellent ≥ 0.75 (Cicchetti & Sparrow, 1981).

### Fingerprinting index

Fingerprinting is described in depth by Finn et al. (2015). For each functional connectivity matrix at timepoint 1, Pearson correlation coefficients are calculated with all connectomes at timepoint 2. The proportion of times a subject’s baseline scan is most correlated with their own scan at the second timepoint (compared to all other scans at that timepoint) is the fingerprinting index for that sample. The correlations are repeated between timepoint 2 scans with every timepoint 1 scan and the two values are averaged to get the overall fingerprinting accuracy. We also calculated group consistency and differential power using scripts from Horien et al. (2018). Group consistency identifies connections that were the least useful in fingerprinting, whereas differential power identifies those that were most useful (for derivations, see Finn et al., 2015).

### Discriminability

Discriminability is a nonparametric multivariate reliability metric conceived by Bridgeford et al. (2021). We calculated discriminability and ran corresponding statistical tests using the *MGC* package in R (Bridgeford et al., 2020). All between-measurement Euclidean distances are computed to generate a distance matrix. To calculate the discriminability of functional connectomes, we treat each connection between two nodes (the correlation values that make up the functional connectome) as a measurement. Thus, for 88 122×122 connectivity matrices, the resulting distance matrix is 10,736×10,736 distances. Discriminability is then calculated as the proportion of within-subject distances that are smaller than between-subject distances. Wang et al. (2020) derive a formula representing the relationship between discriminability and ICC:

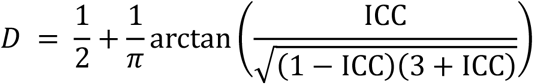

### Edge-level and individual-level reliability associations

To determine if the reliability of functional connections were related to groupwise differences in functional connectivity, we correlated the edge-level effect size of each edge with a continuous edge-level measurement of each reliability metric. To generate the effect sizes, we computed the edgewise Cohen’s *d* effect size of depressed adolescents -healthy volunteers at baseline and at one year. We generated continuous edge-level forms of the three reliability metrics as follows: *ICC* – Mean bootstrapped ICC for each edge; *Fingerprinting* – Differential power and group consistency values for each edge; and *Discriminability* – Mean between-edge distance for each edge.

We also compared individual-level measures to the clinical questionnaires administered at baseline and after one year. We derived individual-level continuous measures of reliability from the original metrics as follows: *ICC* – within subject variance for each individual, *Fingerprinting* – ratio of mean correlation with one’s own connectome to mean correlation with others’ connectomes, and *Discriminability* – Mean between-edge distance for each individual. We compared the continuous reliability measures derived from each metric to both the mean value between the two visits and the change in value across visits. Similarly, medication status was categorized as a binary value (taking/not taking) for psychiatric medications and nonpsychiatric medications. To examine the effect of changing medications on reliability, we compared the change in medication status to the continuous measures. As head motion has a significant impact on reliability, we correlated the measures with average motion and average max framewise displacement. We also explored possible associations between participant age and different reliability measures.

## RESULTS

### Univariate reliability

Edgewise reliability of functional connectivity was poor for both depressed individuals and healthy volunteers [Fig. 1]. However, the depressed participants had higher mean ICC, indicating greater reliability (0.34 > 0.24; Wilcoxon rank-sum: *p* < .001). 1000-fold bootstrapped values were nearly identical to full-sample estimates. Connections to the frontoparietal network and visual association areas were the most reliable in depressed connectomes compared to healthy volunteers.

**Figure 1:**
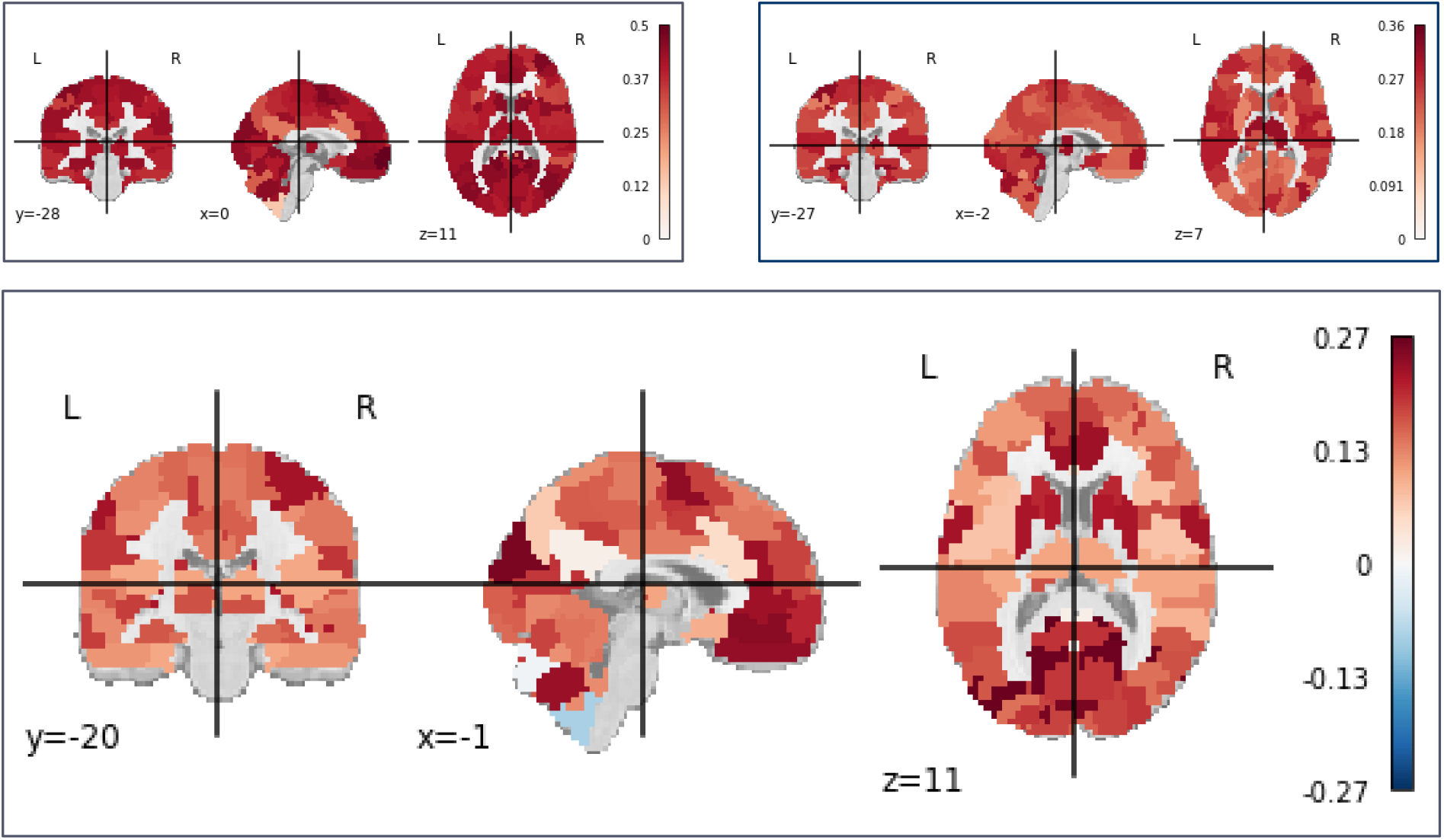
Mean edge-level ICC values for MDD (A) and HV (B), and mean contrast MDD - HV (C). Edge values averaged by ROI.

### Multivariate reliability

Multivariate features of functional connectivity in both groups were reliable [Fig. 2]. Fingerprinting values were greater than chance as estimated by a Poisson (1) distribution (FI_HV_ = 0.53; FI_MDD_ = 0.45; *p* < .001) (Wang et al., 2021). Fingerprinting accuracy did not differ between groups, *X*^*2*^ (1, *N* = 88) = 0.49, *p* = .49. Connections to a region in the prefrontal cortex had the highest group consistency in both healthy and depressed participants, indicating that they reduced identifiability in all subjects [Fig. 3]. These edges had roughly average univariate reliability (μ = 0.27), and group consistency was not correlated with ICC (Pearson’s *r* = 0.11, *p* < .001). This reflects the different reliability aspects captured by these metrics: Group consistency captures edges that are consistent across subjects, i.e., have low between-subject variance, and thus these edges may be reliable at a univariate level. By contrast, the differential power analysis indicated that a diverse array of connections across the brain drove identifiability.

**Figure 2:**
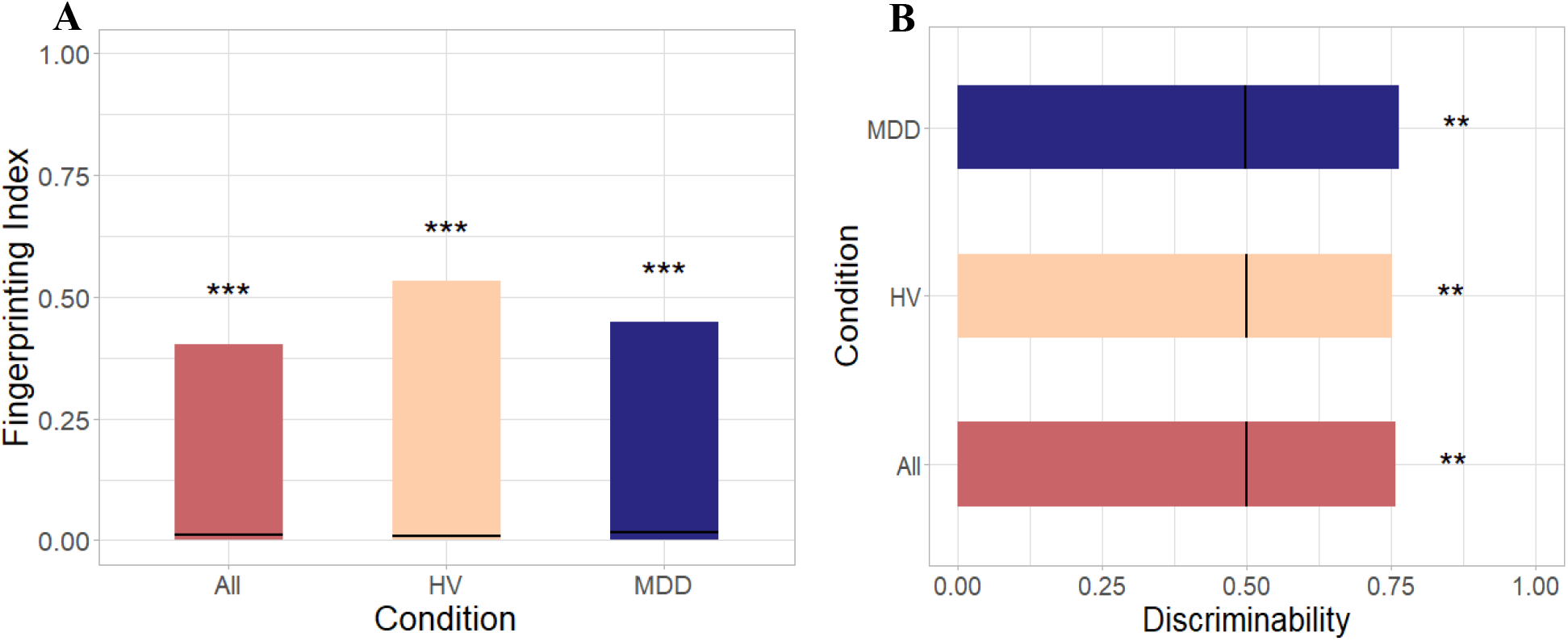
Fingerprinting (A) and discriminability (B) across groups. Black lines indicate chance fingerprinting and discriminability. **: *p* < .01, ***: *p* < .001.

**Figure 3:**
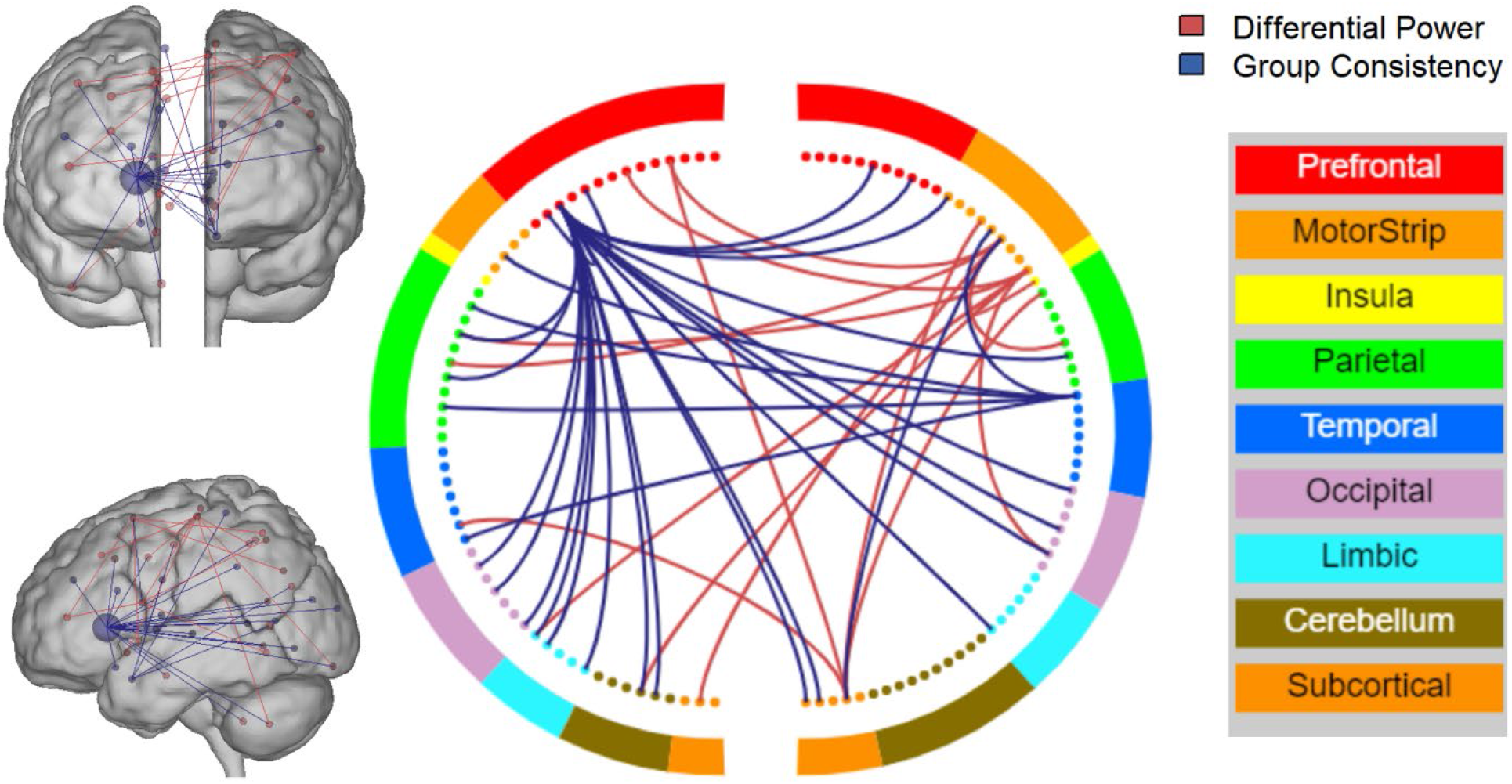
Edges that most improved (red; differential power) and least improved (blue; group consistency) fingerprinting identification. All edges *p* < .005, node degree > 4.

Both groups were more discriminable than chance (*Discr*_HV_ = 0.75; *Discr*_MDD_ = 0.76; 500-fold permutation test *p* < .01) [Fig. 2B]. Discriminability has a deterministic relationship with ICC, which means that an approximately equivalent ICC(2,1) value can be computed for discriminability values (Wang et al., 2020). The overall *Discr*. of 0.76 corresponds to an ICC of 0.83, suggesting that the reliability of multivariate features is much greater than at the univariate level. To put this further into context, we can calculate equivalent discriminability values for the observed univariate ICCs. The overall mean ICC of 0.31 is equivalent to a *Discr*. of 0.56, only just above chance discriminability.

### Association of test-retest reliability with behavioral and clinical measures

The reliability of edges was not associated with the edge-level effect size of group differences between depressed and healthy participants. There was no correlation between ICC, group consistency, differential power, or distance rank [Fig. 4] with MDD-HV effect size at baseline or one year.

**Figure 4:**
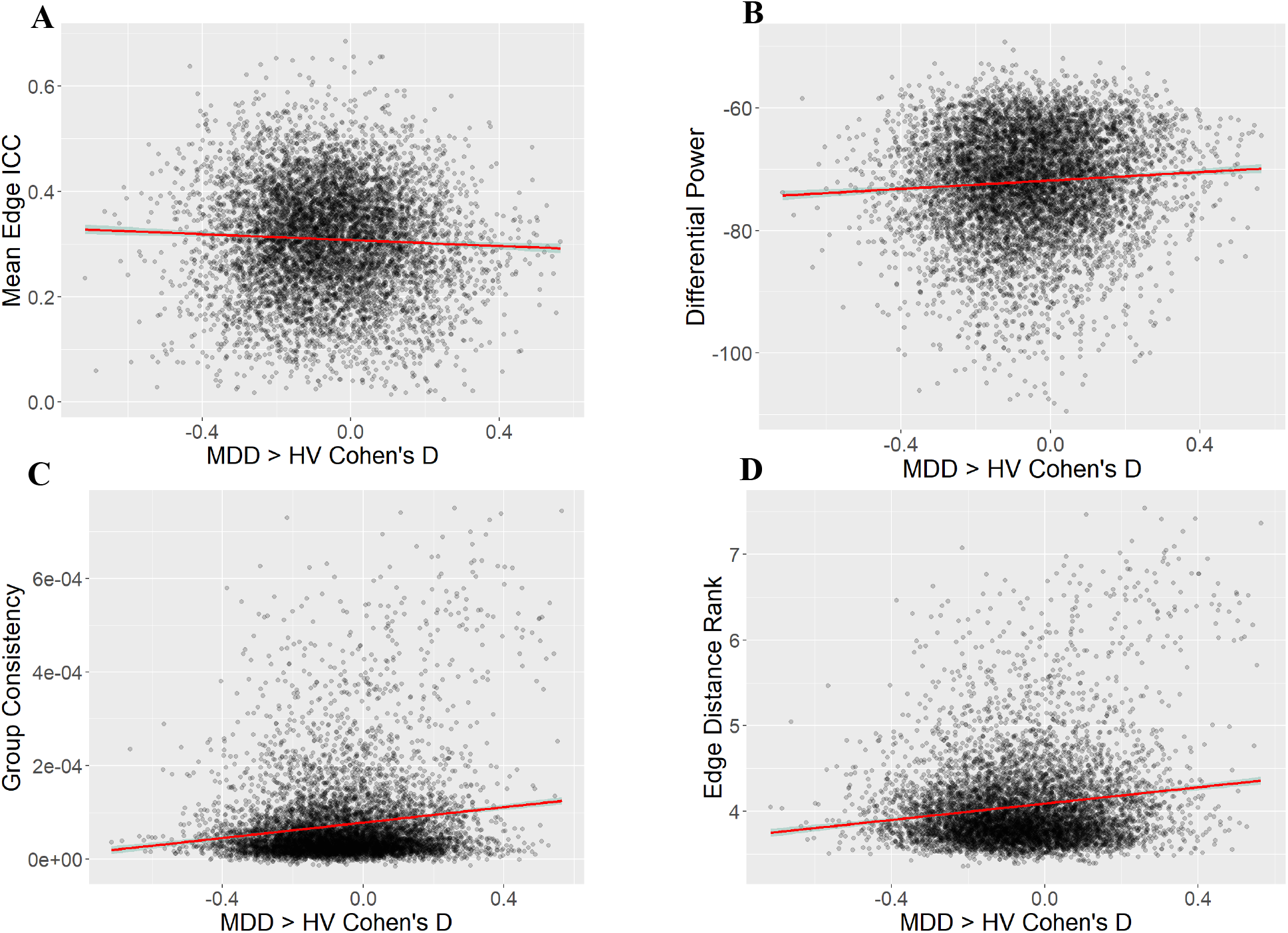
Edge-level ICC (A), Differential Power (B), Group Consistency (C), and Distance Rank (D) correlated with MDD-HV Cohen’s *d* effect size.

Continuous measures of reliability were not correlated with the change or between-session mean of the MFQ, SHAPS, ARI 1 week, or SCARED, nor did they correlate with age or medication status. The continuous measure of fingerprinting was negatively correlated with maximum framewise displacement (Spearman’s *r* = -.38, *p* < .05) and between-session mean framewise displacement (*r* = -.63, *p* < .05) after correction for multiple comparisons, reflecting the sensitivity of fingerprinting to motion and other sources of noise [Fig. 5].

**Figure 5:**
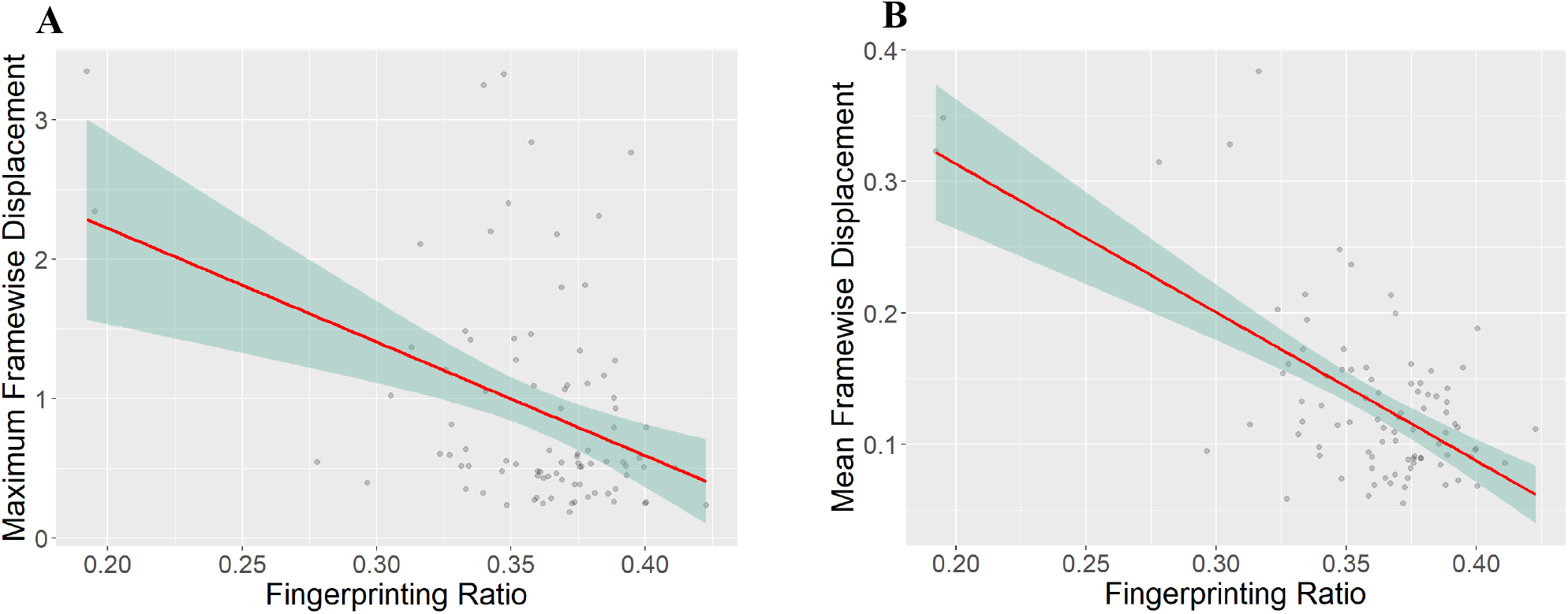
Spearman correlation between continuous measure of fingerprinting and max framewise displacement (A) and between-session mean framewise displacement (B)

## DISCUSSION

The reliability of resting state functional connectivity and what it means for the clinical utility of fMRI is an open line of inquiry. We sought to explore several critical questions in this domain through a multifaceted reliability analysis of a clinically relevant population, namely, depressed adolescents. Over a one-year period, adolescents with depression were more reliable at the edge level than healthy participants, though both groups had poor reliability. Both groups were similarly reliable at the multivariate connectome level. Reliability was not associated with any clinical symptoms.

### Functional connectomes in depressed adolescents are as reliable as in healthy adolescents

Contrary to our hypothesis, depressed adolescents were not less reliable than their healthy peers. At the univariate level, functional connectivity was more reliable in depressed adolescents. However, we did not find strong evidence for an association between depression and reliability. Although the depressed group had higher ICCs on average, both groups were within the expected range of “poor” univariate reliability (<0.4). There were no significant differences in multivariate reliability, with both groups having high discriminability and fingerprinting above chance.

The observed ICC values provide further evidence for the low univariate reliability of fMRI functional connectivity (Noble et al., 2019). While shorter interscan intervals, task-based paradigms, and analyses restricted to cortical nodes can improve reliability, these options offer limited improvement and may not be feasible (Noble et al., 2017, 2021). Increased reliability does not necessarily indicate increased validity, but low reliability places a ceiling on observable effect sizes, increasing type II error and restricting validity.

Although previous studies (Blautzik et al., 2013; Manoach et al., 2001) have found evidence that clinical populations are less reliable, these were conducted with task-based designs in adults with illnesses with cognitive impairment (amnesiatic mild cognitive impairment and schizophrenia). Their sample sizes were also less than 15 participants per group, limiting generalizability. Further reliability analyses in larger and more diverse clinical populations will determine if our findings generalize to other psychiatric illnesses and age groups.

### Both groups have poor univariate reliability but high multivariate reliability

Our findings are consistent with previous work that has found resting state connectivity to be unreliable at the edge level. A review by Noble et al. (2019) found the average ICC of resting state reliability to be 0.29 (95% CI = 0.23-0.36) across 25 studies. Both depressed and healthy adolescents had ICCs within this range, further underscoring the poor reliability of resting state functional connectivity. However, we did find depressed adolescents to be more reliable than their healthy counterparts, with the greatest differences occurring in the frontoparietal and visual association networks. This could be the result of increased correlated noise despite corrections for motion and physiological noise in the preprocessing pipeline. However, we did not observe an association between motion and ICC. There is some evidence that the executive function impairments associated with depression begin in adolescence, which could suggest delayed frontal cortex development (Vilgis et al., 2015). This would result in decreased within-subject variance, potentially improving reliability. Finally, this could be due to decreased between-subject variation in the depressed sample. Although depression is very heterogenous, it could still be the case that a group of depressed participants are more similar to each other than a healthy sample. With within-subject variance constant, decreased between-subject variance would result in increased reliability.

Between-group differences in reliability were only present at the univariate level. Both groups had high multivariate reliability. Directly comparing fingerprinting across populations is difficult due to substantial impacts from sample size, scan length, and age (Horien et al., 2018; Waller et al., 2017). However, a meta-analysis of four datasets by Horien et al. (2019) found similar fingerprinting index values in an adolescent dataset of a similar size. Fingerprinting is noted to be sensitive to motion and other sources of noise (Bridgeford et al., 2021; Horien et al., 2019). In line with this, we observed a negative association between motion and fingerprinting.

Discriminability suggested high reliability, with equivalent ICCs in the “excellent” range. This supports the claim that discriminability is robust to noise (Bridgeford et al., 2021). Further study is needed to determine if high discriminability is indeed reflective of increased validity, i.e., the ability to observe a ground truth association. However, we find support for discriminability as a multivariate reliability metric that captures a level of stability within the connectome. The high multivariate reliability observed here supports previous recommendations that shifting towards multivariate methods can improve reliability (Finn & Rosenberg, 2021; Kragel et al., 2021; Marek et al., 2022; Tetereva et al., 2022). Multivariate reliability in functional connectivity may be higher due to high-dimensionality variance structures in the connectome. Whereas a single edge may be unreliable, zooming out and looking at the larger picture can reveal stable patterns. While some multivariate analyses require additional samples for hyperparameter tuning or to balance the large number parameters, less complex models can provide adequate power with fewer subjects than univariate analysis. And while multivariate analysis may reduce interpretability or effect localization, this consequence is modest compared to the increase in power (Noble et al., 2022).

### Reliability is not associated with behavior at the edge or individual level

We observed no association between edge reliability from any of the metrics and between-group effect size. This supports previous findings that more reliable edges do not have greater utility – that is, they are not better for prediction or observations of brain-behavior associations (Noble et al., 2017). Similarly, we found no association between any of the clinical measures and reliability at the individual level. This underscores the distinction between reliability and validity, especially at the edge level. While edges with low reliability would obscure any true effects, those with high reliability are not necessarily more useful. Noise from sources like head motion or respiration rates can be reliable, and some processing methods like motion regression decrease reliability while increasing the chance of observing a true effect (Noble et al., 2017). Similarly, an individual’s reliability was not associated with their symptoms, even for ICC, where there was a between-group difference. Optimizing reliability is a balancing act that must be done with careful regard for the goal of validity. This motivates the design and testing of new reliability metrics that are tied to validity.

### Limitations

As this was an exploratory investigation, further study is needed to determine if these results replicate in larger and more diverse depressed populations. Our sample was relatively small and predominantly consisted of white, high socioeconomic status (SES) youths from the DC-Maryland-Virginia area. Comparing reliability metrics is difficult – this study is a first step in interpreting the perspectives given by a range of approaches. In investigating the relationship between reliability and validity, we used between-group effect size as a rough proxy for edgewise brain-behavior associations. This was not intended to be a thorough analysis of differences in functional connectivity between depressed and healthy adolescents; rather, we sought to determine if more reliable edges were more likely to have larger effect sizes. Similarly, continuous measures derived from our reliability metrics are not directly comparable to the metrics themselves. These were also intended to reflect relative individual or edgewise reliability. We performed Spearman’s correlations to account for this, which only consider monotonic rank correlations rather than scaled. Comparisons of multivariate and univariate reliability are approximate, as they represent different dimensions of variance. Multivariate and univariate analyses have unique conditions and requirements, and reliability can affect the results of these analyses quite differently. We converted discriminability values to ICC values in order to contextualize the values from a new metric, but these conversions should be viewed as experimental.

## CONCLUSIONS

We characterized the test-retest reliability of rsfMRI functional connectivity in 88 adolescents with and without major depressive disorder. Both depressed and healthy adolescents had low univariate reliability but high multivariate reliability, supporting the increasing shift towards multivariate models in the search for reproducible brain-behavior associations. Reliability was not associated with any clinical measures, although depressed adolescents had higher univariate reliability than healthy volunteers. Overall, we found that individuals with major depressive disorder were no less reliable than healthy participants, suggesting that approaches to optimize reliability and improve biomarker identification may generalize well to this population.

## Data Availability

All data and code referenced in the present study will be made available after an embargo period.

